# Molecular hydrogen for outpatients with Covid-19 (Hydro-Covid): a phase 3, randomised, triple-blinded, adaptive, placebo-controlled, multicentre trial

**DOI:** 10.1101/2024.02.23.24303304

**Authors:** Yoann Gaboreau, Aleksandra Milovančev, Carole Rolland, Claire Eychenne, Jean-Pierre Alcaraz, Cordelia Ihl, Roseline Mazet, François Boucher, Celine Vermorel, Sergej M. Ostojic, Jean Christian Borel, Philippe Cinquin, Jean-Luc Bosson, the HydroCovid Investigators

## Abstract

**Background:** Due to its antioxidative, anti-inflammatory, anti-apoptosis, and antifatigue properties, molecular hydrogen (H_2_) is potentially a novel therapeutic gas for acute coronavirus disease 2019 (COVID-19) patients.

**Aim:** To determine the efficacy and safety profile of hydrogen rich water (HRW) to reduce the risk of progression of COVID-19.

**Design and settings:** We conducted a phase 3, triple-blind, randomized, placebo-controlled trial to evaluate treatment with HRW started within 5 days after the onset of signs or symptoms in primary care patients with mild-to-moderate, laboratory-confirmed COVID-19 and at least one risk factor for severe COVID-19 illness.

**Method:** Participants were randomly assigned to receive HRW or placebo twice daily for 21 days. The composite primary endpoint was the incidence of clinical worsening (dyspnea, fatigue) associated with a need for oxygen therapy, hospitalization or death at day-14; the incidence of adverse events was the primary safety end point.

**Results:** A total of 675 participants were followed up until day-30. 337 in the HRW group and 338 in the placebo group. Baseline characteristics were similar in the two groups. HRW was not superior to placebo in preventing clinical worsening at day-14: in H_2_ group, 46.1% met a clinical deterioration, 43.5% in the placebo group, Hazard Ratio 1.09, 90% confidence interval [0.90-1.31]. One death was reported in the H_2_ group and 2 in the placebo group at day-30. Adverse events were reported in 91 (27%) and 89 (26.2%) participants respectively.

**Conclusion:** Twice-daily ingestion of HRW from the onset of COVID-19 symptoms for 21 days did not reduce clinical worsening.

**How this fits in:**

- Only a few molecules specially developed against SARS-CoV-2 can limit impact of COVID-19 (vaccines, monoclonal antibodies or antiviral drugs)
- Using their multiple properties, H2 may play a key role in preventing the severe and post-acute forms of COVID-19
- Taking twice daily Hydrogen Rich Water (HRW) was not efficacious to prevent severe COVID-19 in at risk COVID-19 patients.
- HRW confirmed a very safe profil

## INTRODUCTION

To date, more than 800 million cases of coronavirus disease 2019 (COVID-19) were confirmed, with a remarkable growing up with Omicron variant during December 2021. Over 7 million deaths worldwide were reported caused by severe acute respiratory syndrome coronavirus 2 (SARS-CoV-2).(1) A slight percentage of COVID-19 patients needed hospitalization, predominantly older adults and persons with pre-existing conditions (e.g., obesity, diabetes mellitus, hypertension or disabilities conditions).(2–4) To avoid these severe complications, several vaccines were developed in record time. These are highly effective in reducing COVID-19-related hospitalizations, admissions to the intensive care unit and deaths.(5) Although two thirds of the world’s population was vaccinated with at least one dose, one third with a booster dose by early December 2023, vaccine coverage still appears to be too sparse and access is uneven.(6) In the same time, antiviral therapies for reducing the risk of severe COVID-19 are emerging. They have been approved at the end of 2021 and a few countries are introducing molnupinavir or nirmatrelvir/ritonavir in their therapeutic arsenal.(7,8) Their implantation in primary care still faces several obstacles: availability in different countries, access to populations, restrictions on their use, conditions of use during the first days of the disease, and a cost-effectiveness ratio that remains to be demonstrated. Other therapeutic approaches have been explored, such as the reuse of molecules like chloroquine, ivermectin, doxycycline, azythromycin, colchicine, or vitamin D. All of these strategies did not reduce the risk of developing a severe form of COVID-19 in the primary care setting.(9–15) Inhaled budesonide might provide a small benefit in high-risk patients, but only one serious study suggests this, and these data need to be confirmed in a larger population.(16) Molecular hydrogen (H_2_) represents a novel approach. Potential preventive and therapeutic applications of H_2_ in various acute and chronic clinical conditions are strongly suggested.(17–20) Thus, H_2_ could be a possible adjunct therapy especially in COVID-19, to combat an excessive proinflammatory response, and in particular increased oxidative stress and apoptosis, due to its anti-inflammatory anti anti-oxidative properties.(17,21–24) Recent studies have confirmed the benefit of H_2_/O_2_ mixing in severe COVID-19 to limit complications, or in combination with a rehabilitation programme in post-acute COVID-19.(25–27) By far the easiest and most practical method of H_2_ administration is oral ingestion of Hydrogen Rich Water (HRW), which has been widely used with a very low rate of side effects. (18,28–30) Moreover, the US-FDA considered HRW as GRAS (Generally Recognized As Safe). Since H_2_ has been shown to be effective on pro-inflammatory agents, it could reduce the destructive cytokine storm caused by SARS-CoV-2 at an early stage.(31) We hypothesize that the administration of HRW during the very first days of the disease, at the stage of mild or moderate ambulatory COVID-19, can avoid the inflammatory cascade leading to the cytokine storm and the dramatic consequences of severe COVID-19. Maintaining H_2_ intake during the 3 weeks of COVID-19 could limit the disabling symptoms present in acute phase, such as dyspnoea and fatigue, and progression to severe symptoms.

## Material and methods

### Study design

A phase 3, double-blinded, parallel-group, randomised, placebo-controlled trial (RCT) evaluating the safety and efficacy of H_2_ for COVID-19 disease in adult outpatients was initiated on January, 2021.

This trial was conducted in 5 French and 1 Serbian regions. It was coordinated by the TIMC public laboratory (Grenoble, France). The trial was done in accordance with the principles of the International Conference on Harmonisation of Good Clinical Practice guidelines.

An inclusion visit by video teleconsultation, or, alternatively, by telephone with a trained physician investigator was arranged in France. In Serbia, inclusion and product delivery were carried out directly by the investigators during the medical visit at general practice doctor’s office. Socio-demographic characteristics and comorbidities were collected at baseline. Data were collected every day during the first month on a paper CRF at home by participants. The primary outcome was collected at Day 12-14 by video teleconsultation, phone or during medical visit by physician investigators according to the health organization of the 2 countries. Secondary outcomes were collected at 1, 3, and 12 months by postal questionnaire and phone calling by research team. All adverse events were reported.

### Participants

Outpatients with mild biological confirmed COVID-19, according to World Health Organization (WHO) guidelines, and with at least one risk factor, were eligible, regardless of whether they had been vaccinated against SARS-CoV-2. Inclusion and exclusion criteria were detailed in supplementary material.

### Randomisation and masking

Eligible and consenting subjects were then randomized in a double-blind fashion to either intervention (HRW) or placebo group by computer-generated random numbers in an 1:1 ratio. Randomisation was stratified in blocks of four stratified by age (< 70 or ≥ 70 years). Both the HRW pill and the placebo were effervescent pills packaged in identical shaped bottles, thus maintaining blinding procedure. The trial team, investigators and participants are not informed of treatment allocation until all participants have completed the one-year follow-up visit.

### Interventions

All participants received usual standard of care for COVID-19 provided by their general practitioners. High concentration HRW was prepared via H_2_-producing tablets (Drink HRW). Participants consumed 1 tablet twice daily in 250 mL of water. Placebo contained identical ingredients to the hydrogen supplement, but instead of metallic magnesium the placebo contained magnesium carbonate.

### Outcomes

The primary endpoint is a composite endpoint of symptom worsening (dyspnoea and fatigue), O2 loading at home or in emergency room, hospitalization (not only a need for the emergency service) and death occurring within 14 days of inclusion in the study. Secondary outcomes included time to clinical improvement, number of days with dyspnoea or fatigue, time to hospitalization for any cause or due to COVID-19 progression, all-cause mortality and time to death from any cause, quality of life and quality of sleep, adverse reactions to the study medications, and the proportion of participants who are non-adherent with the study. All secondary outcomes were assessed up to one month following randomization.

### Safety

Safety endpoints included adverse events occurring during the treatment period (from day 28 or earlier), serious adverse events, and adverse events resulting in discontinuation of treatment or placebo.

### Statistical analysis

Quantitative variables were described as median and quartiles, qualitative variables were described as frequencies and percentages. Time-to-event variables were described the same way as qualitative variables.

The primary analysis compared proportions of patients in the two groups measuring the efficacy of H_2_ compared with placebo. It was assessed by the first primary efficacy end point recording up to day-14, using the Kaplan–Meier method to account for all patients, including those prematurely withdrawn from the trial or lost to follow-up. Adjusted hazard ratio was calculated, stratified on age 70, and associated 90 confidence intervals for analysis using Cox proportional model. Results for physical fatigue, mental symptoms, breathlessness and hospitalization/oxygenotherapy/death separately were presented identically.

For secondary outcomes, treatment compliance and persistent symptoms were compared across groups using Fisher’s exact test. Time to resolution of symptoms between groups was analysed by Mann-Whitney test. The EQ5D5L was analysed using a linear mixed effects model to model all times, using the individual as a random effect, and an interaction between the time (discrete) and the randomisation group. In the same way, the PSQI was analysed using a linear mixed effects model. Sub-group analyses were conducted by adding an interaction term between the treatment and the parameter in the Cox model for the main analysis.

The planned enrollment of 700 participants was selected to ensure greater than 95% power to demonstrate superiority in the primary end point at a one-sided 2.5% alpha level if the underlying event rates were 24% with HRW and 32% with placebo. All analyses were conducted with the use of STATA, version 17 (StataCorp, College Station, Texas).

Interim analyses were not planned. We planned re-evaluation of the event rate when clinical worsening could be evaluated for approximately 100 patients. We planned to stop the study if the event rate was too low, increasing the sample size beyond feasibility.

## Results

### Participants

#### Flow chart and Table of population characteristics

Between January 22, 2021 and March 24, 2022, a total of 700 patients were included at baseline and were randomized into the H_2_ group or placebo group. Twenty-five participants were excluded distributed equally between the 2 groups (Figure 1). Finally, 675 patients analyzed at the follow-up period (day 30). Baseline demographic and clinical characteristics were generally similar in the two groups (Table 1). Overall, 69.5% participants had had onset of signs or symptoms 3 days or less before randomization, 71.7% were vaccinated against SARS-CoV-2 and half (53%) were infected by Delta variant, 38% by Omicron. The most common risk factors were age over 60 years (41.6%) and obesity (28.6%). Survival status were confirmed for all at day-30.

**Figure 1.**
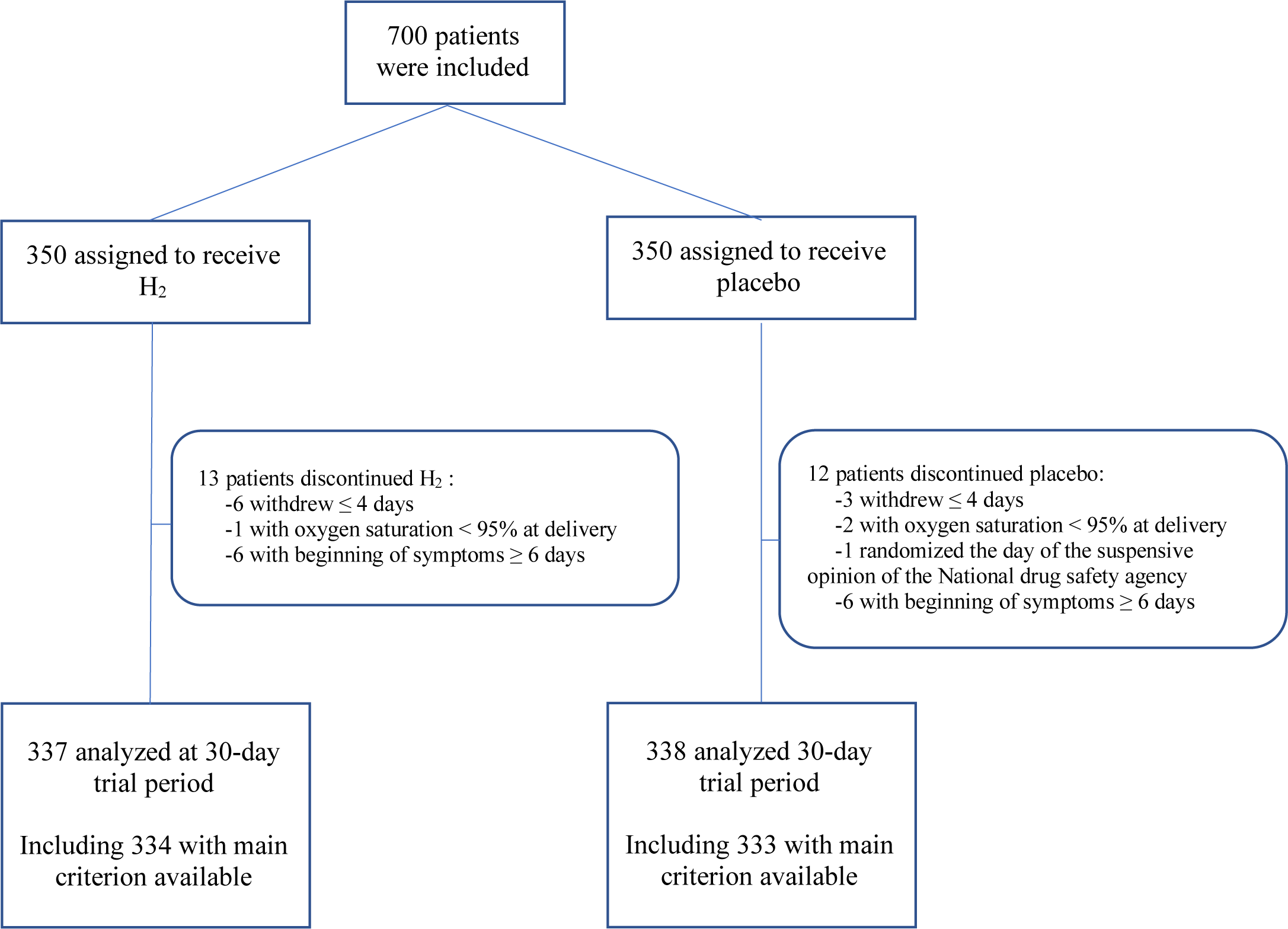
Randomization and Flow of Participants from Baseline.

**Table 1.**
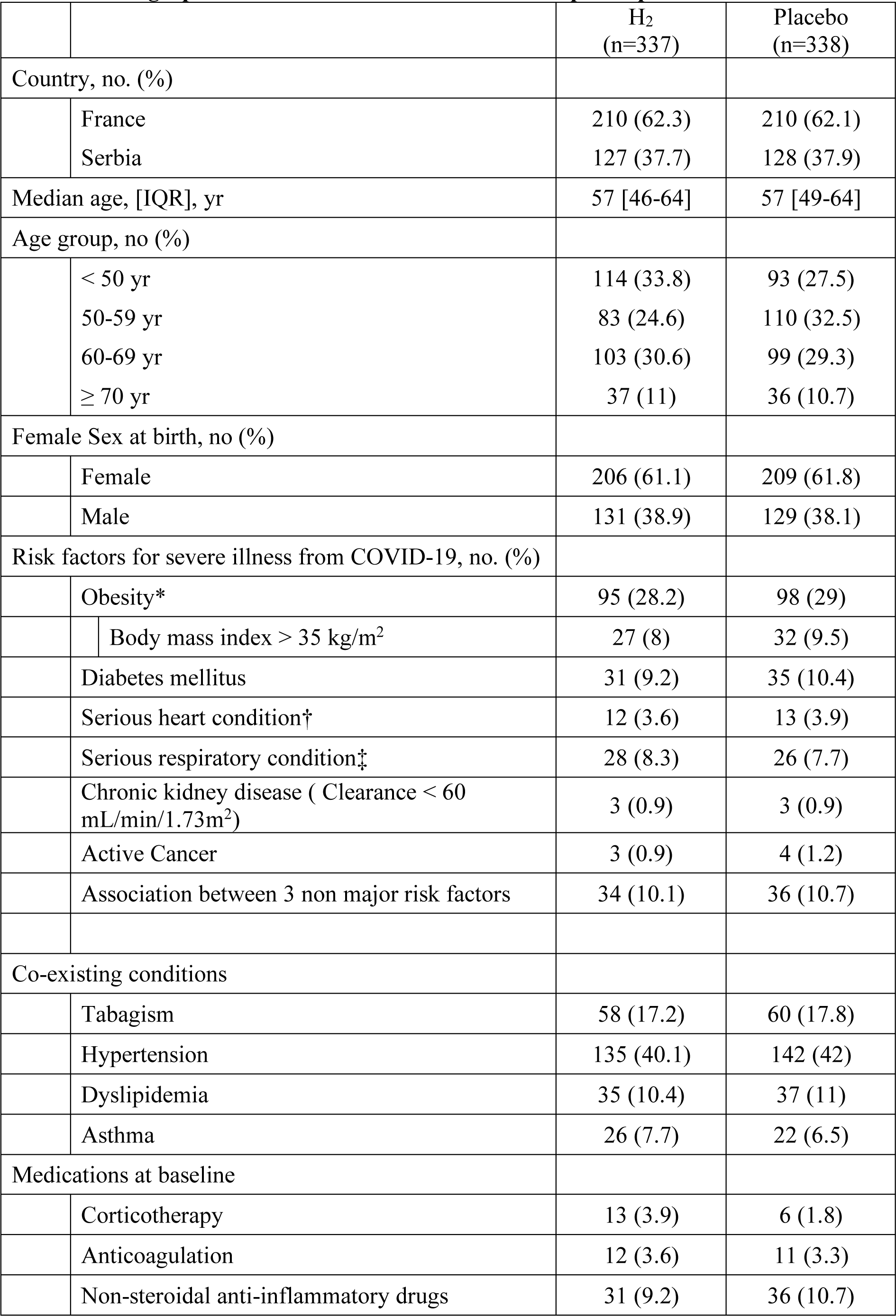
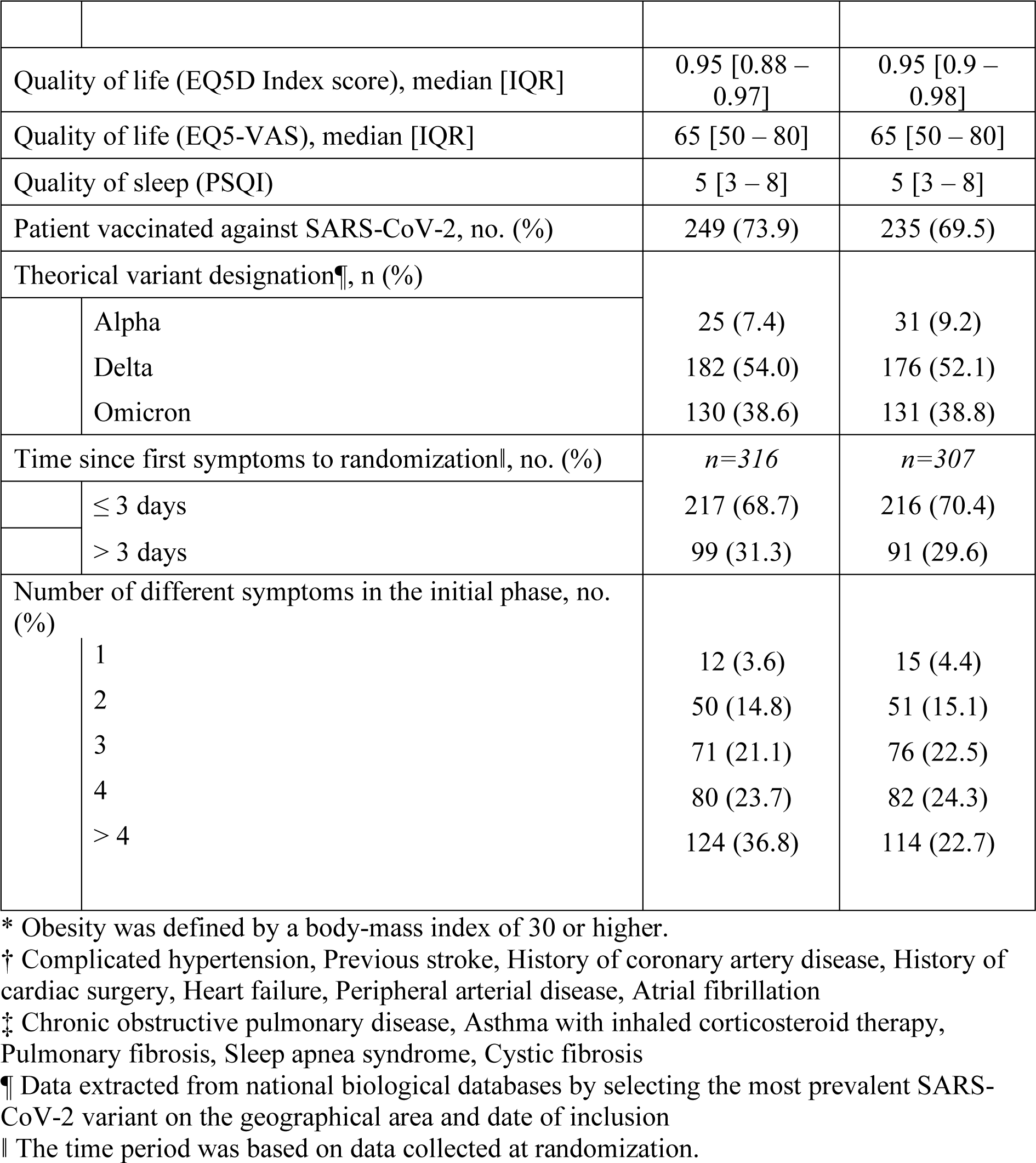
Demographic and clinical characteristics of the participants at baseline.

#### Efficacy

Hydrogen was not superior to placebo at day-14 in preventing clinical worsening. In H_2_ group 154/334 participants (46.1%) experienced a clinical deterioration according to definition of the primary endpoint, 145/333 (43.5%) in the placebo group, Hazard Ratio: 1.09, 90% Confidence Interval [0,90-1.31], p: 0.479 (Graph 1). Among the clinical criteria composing the main composite criterion: physical or mental fatigue, breathlessness, need for oxygen therapy, hospitalization or death, none of them was improved by hydrogen compared with placebo (Table 2). A subgroup analysis was conducted exploring the interaction between the main criterion and age, nature of the SARS-CoV-2 variant, vaccination status or country of inclusion. No statistical interaction was found. A sensibility analysis was conducted excluding the participant when the clinical worsening appeared in the first day after randomization. In this case, 126/306 participants (41.2%) experienced a clinical deterioration in the H_2_ group, 116/304 (38.2%) in the placebo group, p: 0.389.

**Graph1:**
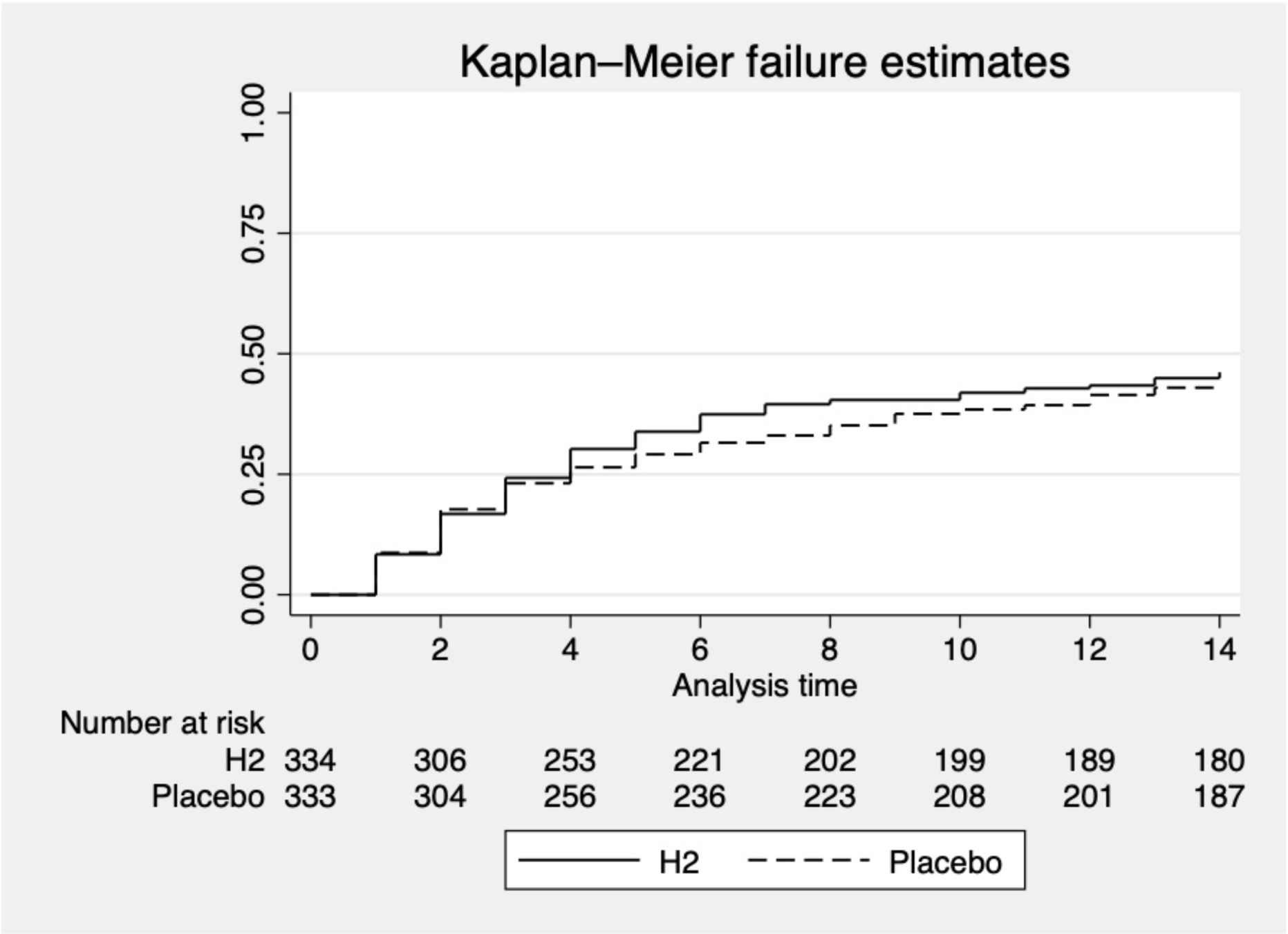
Time-to-Event Analysis of clinical worsening at Day 14. Shown are Kaplan–Meier curves. Clinical worsening is defined as a composite endpoint of symptom worsening (dyspnea and fatigue) at home or in emergency room, hospitalization or death occurring within 14 days of inclusion in the study. Fatigue worsening was defined by a 25% increase via the Chalder scale or via daily visual analog scale (VAS) self-assessment for fatigue. Dyspnea worsening was defined by a 25% increase of the mMRC (modified Medical Research Council) scale or a daily VAS self-assessment

**Table 2:**
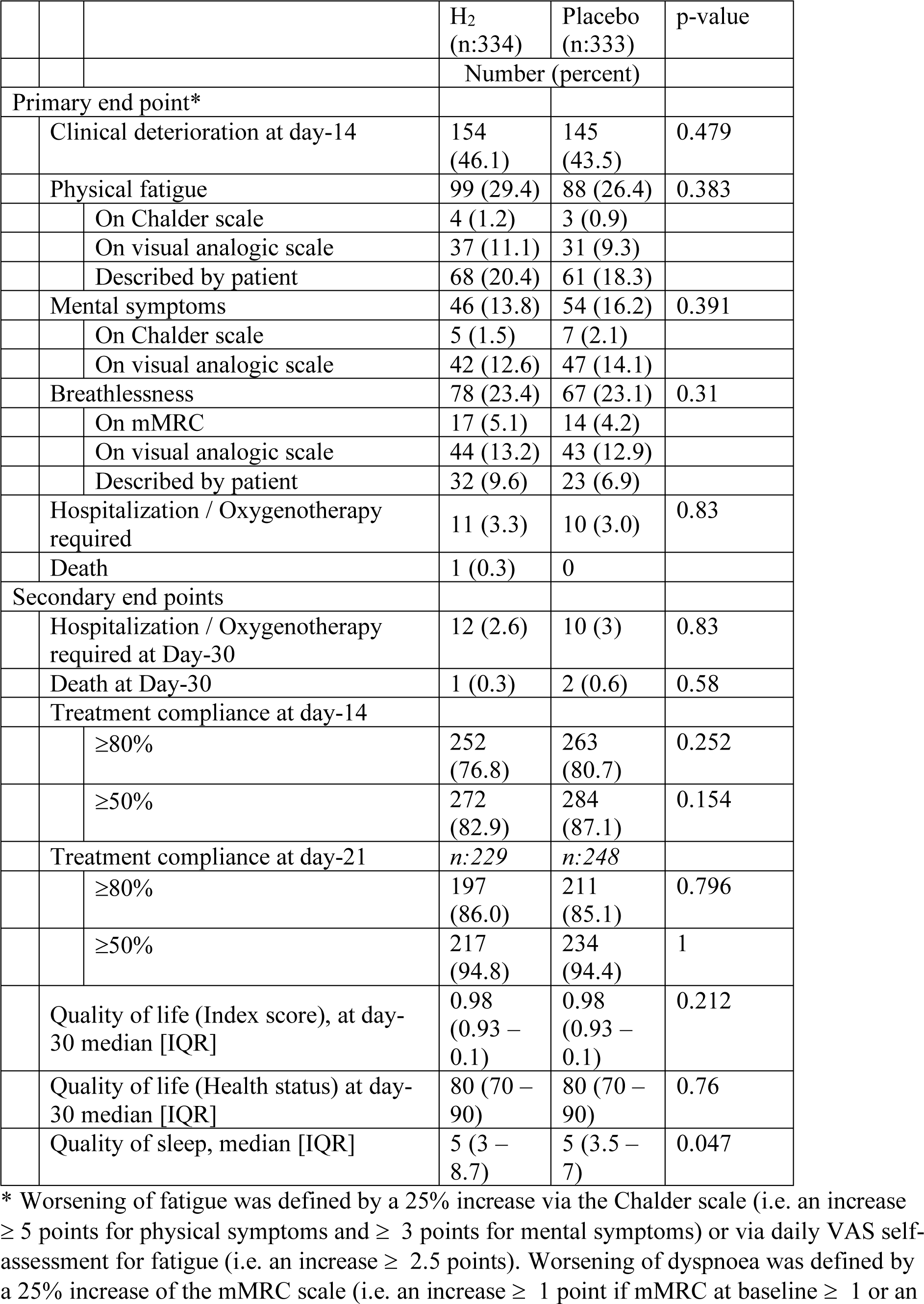
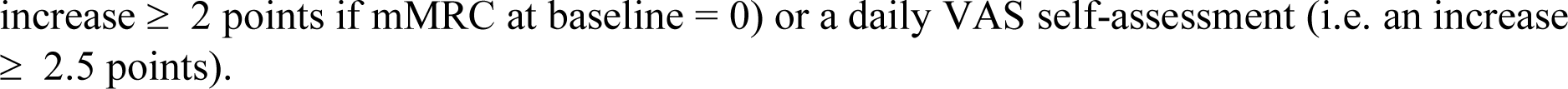
Primary and secondary outcomes.

|  |  |  | H<sub>2</sub><br>(n:334) | Placebo<br>(n:333) | p-value |
| --- | --- | --- | --- | --- | --- |
|  |  |  | Number (percent) |  |  |
| Primary end point* |  |  |  |  |  |
|  | Clinical deterioration at day-14 |  | 154<br>(46.1) | 145<br>(43.5) | 0.479 |
|  | Physical fatigue |  | 99 (29.4) | 88 (26.4) | 0.383 |
|  |  | On Chalder scale | 4 (1.2) | 3 (0.9) |  |
|  |  | On visual analogic scale | 37 (11.1) | 31 (9.3) |  |
|  |  | Described by patient | 68 (20.4) | 61 (18.3) |  |
|  | Mental symptoms |  | 46 (13.8) | 54 (16.2) | 0.391 |
|  |  | On Chalder scale | 5 (1.5) | 7 (2.1) |  |
|  |  | On visual analogic scale | 42 (12.6) | 47 (14.1) |  |
|  | Breathlessness |  | 78 (23.4) | 67 (23.1) | 0.31 |
|  |  | On mMRC | 17 (5.1) | 14 (4.2) |  |
|  |  | On visual analogic scale | 44 (13.2) | 43 (12.9) |  |
|  |  | Described by patient | 32 (9.6) | 23 (6.9) |  |
|  | Hospitalization / Oxygenotherapy required |  | 11 (3.3) | 10 (3.0) | 0.83 |
|  | Death |  | 1 (0.3) | 0 |  |
| Secondary end points |  |  |  |  |  |
|  | Hospitalization / Oxygenotherapy required at Day-30 |  | 12 (2.6) | 10 (3) | 0.83 |
|  | Death at Day-30 |  | 1 (0.3) | 2 (0.6) | 0.58 |
|  | Treatment compliance at day-14 |  |  |  |  |
|  |  | ≥80% | 252<br>(76.8) | 263<br>(80.7) | 0.252 |
|  |  | ≥50% | 272<br>(82.9) | 284<br>(87.1) | 0.154 |
|  | Treatment compliance at day-21 |  | *n*:229 | *n*:248 |  |
|  |  | ≥80% | 197<br>(86.0) | 211<br>(85.1) | 0.796 |
|  |  | ≥50% | 217<br>(94.8) | 234<br>(94.4) | 1 |
|  | Quality of life (Index score), at day-30 median [IQR] |  | 0.98<br>(0.93 – 0.1) | 0.98<br>(0.93 – 0.1) | 0.212 |
|  | Quality of life (Health status) at day-30 median [IQR] |  | 80 (70 – 90) | 80 (70 – 90) | 0.76 |
|  | Quality of sleep, median [IQR] |  | 5 (3 – 8.7) | 5 (3.5 – 7) | 0.047 |

\* Worsening of fatigue was defined by a 25% increase via the Chalder scale (i.e. an increase $\geq 5$ points for physical symptoms and $\geq 3$ points for mental symptoms) or via daily VAS self-assessment for fatigue (i.e. an increase $\geq 2.5$ points). Worsening of dyspnoea was defined by a 25% increase of the mMRC scale (i.e. an increase $\geq 1$ point if mMRC at baseline $\geq 1$ or an
increase $\geq 2$ points if mMRC at baseline = 0) or a daily VAS self-assessment (i.e. an increase $\geq 2.5$ points).

The large majority of hospitalizations occurred during the first 2 weeks, and all 3 deaths were attributed to respiratory distress secondary to COVID-19. Quality of life improved during the first month of follow-up (relative difference of 20.7%), but strictly in parallel between the 2 groups. The time to resolution of symptoms was 11 days, [IQR: 7-17] in the H_2_ group and 14 days [IQR: 7 - 20] in the placebo group, p: 0.186. At one month of follow-up, 39% of participants had persistent symptoms with no difference between the two groups: anosmia, dysgeusia for 58 (10%), breathing difficulties for 55 (9.5%), cough for 62 (10.7%), arthralgia for 65 (10.6%).

#### Safety

The incidence of adverse events that emerged during or after the treatment period was similar among the two groups (Table 3). None of the serious adverse events (SAE) graded 5 or 4 related to drug or placebo were reported during the acute phase. Grade 3 events were more frequent, and 45 of the 48 serious situations were linked to the severity of COVID-19 and hospitalization. Suspected serious unexpected adverse reaction was found in a single patient after 3 days of treatment: boudless energy, insomnia, visual and hearing hallucination, anxiety, lower limb oedema. These symptoms were attributed to placebo group.

**Table 3.** Summary of Adverse Events, Serious Adverse Events, and Adverse Events Leading to Discontinuation through Day 30.*

| **Adverse Event Category** | **H<sub>2</sub>**<br>**(N=337)** | **Placebo**<br>**(N=338)** |
| --- | --- | --- |
| **Events that emerged during treatment period** |  |  |
| Patients with adverse events — no. (%) | 91 (27) | 89 (26.2) |
| Any adverse event | 126 (37.4) | 124 (36.7) |
| Serious adverse event | 21 (6.2) | 27 (8) |
| Maximum grade 3 or 4 adverse event | 7 (2.1) | 11 (3.3) |
| Maximum grade 5 adverse event | 1 | 2 |
| Discontinued drug or placebo because of adverse event | 16 (4.7) | 14 (4.1) |
| Had dose reduction or temporary discontinuation owing to adverse event | 2 | 3 |
| **Events considered to be related to drug or placebo** |  |  |
| Patients with adverse events — no. (%) | 60 (17.8) | 59 (17.5) |
| Any adverse event | 76 (22.6) | 74 (21.9) |
| Serious adverse event | 1 | 0 |
| Maximum grade 3 or 4 adverse event | 1 | 0 |
| Maximum grade 5 adverse event | 0 | 0 |
| Discontinued drug or placebo because of adverse event | 4 (1.2) | 5 (1.5) |
| Had dose reduction or temporary discontinuation owing to adverse event | 23 (6.8) | 21 (6.2) |

\*Shown are data for all patients who received at least one dose of drug or placebo
†All reported deaths were related to COVID-19; causes of death included COVID-19 pneumonia (3 patients).

An ulcerative syndrome and a hepatic colic were detected in the H_2_ group. A drug-drug interaction involving azythromycin and colchicine was detected, requiring hospitalization.

H_2_ treatment was well tolerated, 85.5% of participants completed the 21 days treatment with a good compliance (Table 2). Among the reasons for discontinuation of study treatment, 10 patients described allergic-type reactions (5 in each group), and 92 patients (44 versus 48) experienced expected side effects such as diarrhea or abdominal pain. In a lesser proportion, 16 patients experienced vomiting (8 in each group). One patient reported a sensation of hypertension and was randomized to the placebo group. A few patients declared sensations of metal rubbing on certain dental prostheses only in the H_2_ group. Other Class 1 side effects reported by patients were less than 1%.

## Discussion

### Summary

These data from the Hydro-Covid phase 3 trial in non-hospitalized at-risk adults with COVID-19 indicate that HRW, initiated within 5 days after the onset of symptoms, does not reduce the risk of hospitalization for any cause or death through day-30. The speed of symptoms recovery was equivalent in the 2 arms. All patients had regained a good quality of life and good sleep whatever the treatment at Day-30. The safety profile of HRW was reassuring, with a frequency of AEs in line with expectations.

### Strengths and limitations

The absence of a detected effect may be the result of several factors: a composite primary endpoint that was too “soft”. We opted for a patient centered approach about their symptoms and discomfort measured by VAS and validated scales, combined with stricter criteria such as hospitalization, oxygen therapy and death. This data is illustrated by the fact that 44.8% of patients met the primary endpoint, mainly symptomatic criteria, while 3.3% met severe COVID-19 criteria. The profile of the included patients suggested that they were initially in good health, that they were interested in a dietary supplement as the subject of this trial, and that their exposure to pathologies and risk factors was low. In addition, the environmental context, with the rise of SARS-Cov-2 vaccination, probably played a supplementary protective role. Indeed, public policies have preferentially targeted at-risk populations in order to limit the impact on the hospital system and its associated burden. Patients were recruited for this study during the primary and booster vaccination phases. The protective effect has now been demonstrated, reducing the risk of hospitalization or death in vaccinated patients by a factor of 3 to 10, depending on the number of associated co-morbidities(5,32).

Despite the negative results, and the limitations described above, this study has methodological advantages: it took place solely in patients’ homes, with recruitment exclusively in primary care. In addition, it validated the recruitment of patients into a clinical trial by teleconsultation, accompanied by a home visit by mobile teams made up of members of the research team or private nurses. This shows that therapeutic trials in primary care are feasible. The COVID-19 clinical trials have had a leverage effect in getting research out of hospital centers and proposing organizational innovations to reach out to patients in their own environment. As a case in point, the COVERAGE platform trial involved 213 stakeholders in setting up and running the clinical trial for 1 investigating center(33). Of these, more than half were the mobile teams responsible for the inclusion and home follow-up of the included patients.

### Comparison with existing literature

Other studies have used different concentrations and delivery methods, such as inhalation. Lebaron et al. were able to test changes in biological parameters after 6 months’ exposure to HRW 3 times a day (34). Another trial in 2019 tested the efficacy of ingesting 2 tablets of HRW on physical performance, noting a reduction in heart rate and respiratory rate without any change in VO2 max(35). Mikami et al were able to show the efficacy of taking 500 mL of HRW 30 min before physical effort on an ergometric cycle on VO2max and the Borg scale (36). Botek et al. were able to measure the effect of inhaled H_2_ on effects in terms of improved physical and respiratory function in acute post-COVID-19 patients (27). These data support a biological and clinical effect of H_2_, but the effective dose and conditions of use have yet to be determined.

### Implications for research and/or practice

Molecular hydrogen was recently discovered as a potential therapeutic agent. In three decades, more than 900 clinical trials were described on Pubmed. A thematic issue consisting of 19 review articles on H_2_ medicine was recently published(37). The therapeutic possibilities are numerous, both for sick patients - all the more interesting as the inflammatory component is predominant - and for healthy people. This approach deserves to be brought to the attention of Western research teams, who are poorly represented in this particularly promising field of research.

## Data Availability

All data produced in the present study are available upon reasonable request to the authors

## Additional information

### Ethical aproval

It was approved by the French Agency for the safety of Drugs and Health Products, the Sud Mediterrannée II Committee for the protection of persons (reference 2020 A03137 32) and was compliant with General Data Protection Regulation. The protocol, statistical analysis plan, and additional information are available on clinicaltrials.gov (ID: NCT04716985).

### Competting interests

No author have any conflict of interest with this trial.

## Ackowledgement

The authors would like to sincerely thanks for:

- Helping organisation: LEHMANN, Cabinet infirmier Aurélien CERCEUIL, Cabinet infirmier CHOUGNY-DUQUESNE, Cabinet infirmier LOSA-VILBOUX-BRECHT, Cabinet infirmier Marielle PENZ, Cabinet infirmier PERNEY-LETORT-THUILLIER-PACAUD, Imprimerie des Ecureuils, Robé médical
- Helping communication: BOSSON, Jérome BRAVARD, Morgane BURNEL, Jean-Marie CABRIERES, Déborah CADAT, Leila CHAUMONT, Isabelle CIEREN, Jacques COURCHELLE, Guillaume DEBATY, Bruno DELPEUCH, Norbert DESBIOLLES, Camille EYCHENNE, FLORALIS, Pierre FOURNIER, Carole FREBY, Anne GALLET, Muriel JAKOBIAK, Laboratoire BIOGROUP, Laboratoire BIOMED 21, Laboratoire EUROFINS, Laboratoire ORIADE, Laboratoire UNIBIO, Frédéric LAMBERT, Hervé LELIEVRE, Catherine MANZOTTI, Delphine MARGOT, Séverine MEUNIER, Vincent PEYLE, Simon PERVIER, Philippe PICHON, Guillaume RICHALET, SAMU Isère, Caroline SANCHEZ, Université Inter-Ages du Dauphiné, Catherine ZOPPIS
- Help screening and delivery: ADSC, Mathys ANCEL, Hassan AQRAA, Laura ALBALADEJO, Cassandra BRECVILLE, Alexa COMTE, Mona DAOUD, Camille EYCHENNE, Mathilde FRANCHINO, Maryline GABOREAU, Lila KREBS DROUOT, Aurélien LEFAIVRE, Andreja MAKSIMOVIC, Faustine MONIN, Claire PARADIS, Joris RAGGAZZINI, Cassandra SACCHETTO, Noa SANCHEZ Y PARE, Nada SHOUR, Marijana RANISAVLJEV, Patrik DRID, Nikola TODOROVIĆ, Danijela MUSULIN BANJANIN, Milijana ČELAREVIĆ, Olivera KRAJČINOVIĆ, Tamara BULAJIĆ, Jovana AVAKUMOVIĆ, Biljana KNEŽEVIĆ, Jelena MATEJIĆ,

## Appendix: List of HydroCovid investigators

BARRELIER Marie-Thérèse, BOSSON Jean-Luc, DAVID TCHOUDA Sandra, DESCOMBE Fabrice, GABOREAU Yoann, LANOYE Patrick, MILOVANCEVAleksandra, PARADIS Sabrina, SORS Claire.

## REFERENCES

1. WHO coronavirus (COVID-19) dashboard. Geneva: World Health Organization, 2021 (https://covid19.who.int).

2. Semenzato L, Botton J, Drouin J, Cuenot F, Dray-Spira R, Weill A, et al. Chronic diseases, health conditions and risk of COVID-19-related hospitalization and in-hospital mortality during the first wave of the epidemic in France: a cohort study of 66 million people. Lancet Reg Health Eur. sept 2021;8:100158.

3. Zhou F, Yu T, Du R, Fan G, Liu Y, Liu Z, et al. Clinical course and risk factors for mortality of adult inpatients with COVID-19 in Wuhan, China: a retrospective cohort study. Lancet. 28 mars 2020;395(10229):1054-62.

4. Bosworth ML, Ayoubkhani D, Nafilyan V, Foubert J, Glickman M, Davey C, et al. Deaths involving COVID-19 by self-reported disability status during the first two waves of the COVID-19 pandemic in England: a retrospective, population-based cohort study. Lancet Public Health. nov 2021;6(11):e817–25.

5. Zheng C, Shao W, Chen X, Zhang B, Wang G, Zhang W. Real-world effectiveness of COVID-19 vaccines: a literature review and meta-analysis. Int J Infect Dis. janv 2022;114:252–60.

6. Ritchie H, Mathieu E, Rodés-Guirao L, Appel C, Giattino C, Ortiz-Ospina E, et al. Coronavirus Pandemic (COVID-19). Published online at OurWorldInData.org. Retrieved from: « https://ourworldindata.org/coronavirus » [Online Resource]. 2020;Published online at OurWorldInData.org. Retrieved from: « https://ourworldindata.org/coronavirus » [Online Resource].

7. Owen DR, Allerton CMN, Anderson AS, Aschenbrenner L, Avery M, Berritt S, et al. An oral SARS-CoV-2 M ^pro^ inhibitor clinical candidate for the treatment of COVID-19. Science. 24 déc 2021;374(6575):1586–93.

8. Jayk Bernal A, Gomes da Silva MM, Musungaie DB, Kovalchuk E, Gonzalez A, Delos Reyes V, et al. Molnupiravir for Oral Treatment of Covid-19 in Nonhospitalized Patients. N Engl J Med. 10 févr 2022;386(6):509–20.

9. Butler CC, Dorward J, Yu LM, Gbinigie O, Hayward G, Saville BR, et al. Azithromycin for community treatment of suspected COVID-19 in people at increased risk of an adverse clinical course in the UK (PRINCIPLE): a randomised, controlled, open-label, adaptive platform trial. The Lancet. mars 2021;397(10279):1063–74.

10. Singh B, Ryan H, Kredo T, Chaplin M, Fletcher T. Chloroquine or hydroxychloroquine for prevention and treatment of COVID-19. Cochrane Database Syst Rev. 12 févr 2021;2:CD013587.

11. Tardif JC, Bouabdallaoui N, L’Allier PL, Gaudet D, Shah B, Pillinger MH, et al. Colchicine for community-treated patients with COVID-19 (COLCORONA): a phase 3, randomised, double-blinded, adaptive, placebo-controlled, multicentre trial. The Lancet Respiratory Medicine. août 2021;9(8):924–32.

12. PRINCIPLE Trial Collaborative Group, Dorward J, Yu LM, Hayward G, Saville BR, Gbinigie O, et al. Colchicine for COVID-19 in adults in the community (PRINCIPLE): a randomised, controlled, adaptive platform trial. 23 sept 2021 [cité 17 févr 2022]; Disponible sur: http://medrxiv.org/lookup/doi/10.1101/2021.09.20.21263828

13. Butler CC, Yu LM, Dorward J, Gbinigie O, Hayward G, Saville BR, et al. Doxycycline for community treatment of suspected COVID-19 in people at high risk of adverse outcomes in the UK (PRINCIPLE): a randomised, controlled, open-label, adaptive platform trial. Lancet Respir Med. sept 2021;9(9):1010–20.

14. Popp M, Stegemann M, Metzendorf MI, Gould S, Kranke P, Meybohm P, et al. Ivermectin for preventing and treating COVID-19. Cochrane Database Syst Rev. 28 juill 2021;7:CD015017.

15. Jolliffe DA, Camargo CA, Sluyter JD, Aglipay M, Aloia JF, Ganmaa D, et al. Vitamin D supplementation to prevent acute respiratory infections: a systematic review and meta-analysis of aggregate data from randomised controlled trials. Lancet Diabetes Endocrinol. mai 2021;9(5):276–92.

16. Yu LM, Bafadhel M, Dorward J, Hayward G, Saville BR, Gbinigie O, et al. Inhaled budesonide for COVID-19 in people at high risk of complications in the community in the UK (PRINCIPLE): a randomised, controlled, open-label, adaptive platform trial. The Lancet. sept 2021;398(10303):843–55.

17. Ohta S. Direct Targets and Subsequent Pathways for Molecular Hydrogen to Exert Multiple Functions: Focusing on Interventions in Radical Reactions. Curr Pharm Des. 2021;27(5):595–609.

18. Ohta S. Molecular hydrogen as a preventive and therapeutic medical gas: initiation, development and potential of hydrogen medicine. Pharmacol Ther. oct 2014;144(1):1–11.

19. Ohta S. Molecular Hydrogen as a Novel Antioxidant. In: Methods in Enzymology [Internet]. Elsevier; 2015 [cité 18 févr 2022]. p. 289–317. Disponible sur: https://linkinghub.elsevier.com/retrieve/pii/S0076687914001037

20. Nicolson GL, de Mattos GF, Settineri R, Costa C, Ellithorpe R, Rosenblatt S, et al. Clinical Effects of Hydrogen Administration: From Animal and Human Diseases to Exercise Medicine. IJCM. 2016;07(01):32–76.

21. Russell G, Rehman M, LeBaron T, Veal D, Adukwu E, Hancock J. An Overview of SARS-CoV-2 (COVID-19) Infection and the Importance of Molecular Hydrogen as an Adjunctive Therapy. ROS [Internet]. 2020 [cité 17 févr 2022]; Disponible sur: https://www.rosj.org/index.php/ros/article/view/271

22. Yang F, Yue R, Luo X, Liu R, Huang X. Hydrogen: A Potential New Adjuvant Therapy for COVID-19 Patients. Front Pharmacol. 2020;11:543718.

23. Tw L, Ml M, Sr KH R. A novel functional beverage for COVID-19 and other conditions: Hypothesis and preliminary data, increased blood flow, and wound healing. J Transl Sci [Internet]. 2020 [cité 17 févr 2022];6(6). Disponible sur: https://www.oatext.com/a-novel-functional-beverage-for-covid-19-and-other-conditions-hypothesis-and-preliminary-data-increased%20blood%20flow%20and%20wound%20healing.php

24. Alwazeer D, Liu FFC, Wu XY, LeBaron TW. Combating Oxidative Stress and Inflammation in COVID-19 by Molecular Hydrogen Therapy: Mechanisms and Perspectives. Hasnain MS, éditeur. Oxidative Medicine and Cellular Longevity. 4 oct 2021;2021:1–17.

25. Guan WJ, Wei CH, Chen AL, Sun XC, Guo GY, Zou X, et al. Hydrogen/oxygen mixed gas inhalation improves disease severity and dyspnea in patients with Coronavirus disease 2019 in a recent multicenter, open-label clinical trial. J Thorac Dis. Juin 2020;12(6):3448–52.

26. Shogenova LV, Truong TT, Kryukova NO, Yusupkhodzhaeva KA, Pozdnyakova DD, Kim TG, et al. Hydrogen inhalation in rehabilitation program of the medical staff recovered from COVID-19. Cardiovasc Ther Prev. 16 oct 2021;20(6):2986.

27. Botek M, Krejčí J, Valenta M, McKune A, Sládečková B, Konečný P, et al. Molecular Hydrogen Positively Affects Physical and Respiratory Function in Acute Post-COVID-19 Patients: A New Perspective in Rehabilitation. IJERPH. 10 févr 2022;19(4):1992.

28. Ono H, Nishijima Y, Adachi N, Sakamoto M, Kudo Y, Kaneko K, et al. A basic study on molecular hydrogen (H2) inhalation in acute cerebral ischemia patients for safety check with physiological parameters and measurement of blood H2 level. Med Gas Res. 2012;2(1):21.

29. Fontanari P, Badier M, Guillot Ch, Tomei C, Burnet H, Gardette B, et al. Changes in maximal performance of inspiratory and skeletal muscles during and after the 7.1-MPa Hydra 10 record human dive. European Journal of Applied Physiology. 18 janv 2000;81(4):325–8.

30. Ohta S. Recent Progress Toward Hydrogen Medicine: Potential of Molecular Hydrogen for Preventive and Therapeutic Applications. CPD. 1 juill 2011;17(22):2241–52.

31. Yang L, Xie X, Tu Z, Fu J, Xu D, Zhou Y. The signal pathways and treatment of cytokine storm in COVID-19. Sig Transduct Target Ther. déc 2021;6(1):255.

32. Simard M, Boiteau V, Fortin É, Jean S, Rochette L, Trépanier PL, et al. Impact of chronic comorbidities on hospitalization, intensive care unit admission and death among adult vaccinated and unvaccinated COVID-19 confirmed cases during the Omicron wave. Journal of Multimorbidity and Comorbidity. déc 2023;13:263355652311695.

33. Grenier C, Loniewski M, Plazy M, Onaisi R, Doucet MH, Joseph JP, et al. Implementing an outpatient clinical trial on COVID-19 treatment in an emergency epidemic context: a mixed methods study among operational and research stakeholders within the Coverage trial, Bordeaux (France). Arch Public Health. 3 déc 2022;80(1):245.

34. LeBaron TW, Kura B, Kalocayova B, Tribulova N, Slezak J. A New Approach for the Prevention and Treatment of Cardiovascular Disorders. Molecular Hydrogen Significantly Reduces the Effects of Oxidative Stress. Molecules. 31 mai 2019;24(11):2076.

35. LeBaron TW, Laher I, Kura B, Slezak J. Hydrogen gas: from clinical medicine to an emerging ergogenic molecule for sports athletes 1. Can J Physiol Pharmacol. sept 2019;97(9):797–807.

36. Mikami T, Tano K, Lee H, Lee H, Park J, Ohta F, et al. Drinking hydrogen water enhances endurance and relieves psychometric fatigue: a randomized, double-blind, placebo-controlled study 1. Can J Physiol Pharmacol. sept 2019;97(9):857–62.

37. Ohta S. Development of Hydrogen Medicine and Biology: Potential for Various Applications in Diverse Fields. CPD. févr 2021;27(5):583–4.

